# Measuring Positive Stress Appraisal Among Nursing Students: Development and Psychometric Evaluation of the Nursing Student Positive Stress Scale (NSPSS)

**DOI:** 10.64898/2026.08.30.26361779

**Authors:** Han Yan, Anthony John O’Brien, Seong Hoon Yoon, Toya Shaw, Kat Vakavosaki

**Affiliations:** School of Nursing and Midwifery, The University of Waikato, Private Bag 3105, Hamilton 3240, New Zealand; School of Computing and Mathematical Sciences, The University of Waikato, Private Bag 3105, Hamilton 3240, New Zealand

**Author notes:** Corresponding author, Han Yan, School of Nursing and Midwifery, The University of Waikato, Private Bag 3105, Hamilton 3240, New Zealand.

**Keywords:** Nursing students, Positive stress appraisal, Eustress, Scale development, Psychometric evaluation, Transactional theory of stress and coping

## Abstract

**Background:** Stress research in nursing education has largely focused on distress, stressors, and negative outcomes, although challenging experiences may also support motivation, confidence, learning, and growth when appraised positively.

**Objective:** To develop and evaluate the psychometric properties of the Nursing Student Positive Stress Scale (NSPSS).

**Design:** A methodological instrument development and psychometric evaluation study.

**Methods:** The NSPSS was developed using a deductive, theory-driven approach informed by the transactional theory of stress and coping and positive psychology perspectives. Content validity was assessed by an international nursing expert panel. Psychometric evaluation used national survey data from nursing students in New Zealand. Of 539 responses, 507 were analysed. Exploratory factor analysis (EFA) and confirmatory factor analysis (CFA) were conducted using separate subsamples. Internal consistency was assessed using Cronbach’s alpha and McDonald’s omega, and convergent validity through correlation with Perceived Stress Scale-10 scores.

**Results:** Content validity was strong (I-CVI = .88–1.00; S-CVI/Ave = .975; S-CVI/UA = .800). EFA identified a dominant factor explaining 41.38% of variance (loadings = .528–.735). CFA supported a two-context Academic and Clinical Positive Stress model with correlated residuals between five parallel item pairs, χ²(29) = 60.49, CFI = .970, TLI = .954, RMSEA = .063, SRMR = .065. Internal consistency was good (α = .839; ω = .843). NSPSS scores correlated negatively with PSS-10 scores (r = −.298, p < .001).

**Conclusion:** The NSPSS demonstrated strong content validity, preliminary evidence of structural and convergent validity, and good internal consistency reliability for assessing positive stress appraisal among nursing students. Further validation in independent samples is warranted.

**Impact statement:** The NSPSS provides a theory-driven measure of positive stress appraisal across academic and clinical nursing education contexts, enabling more comprehensive investigation of how nursing students respond adaptively to stress.

**Plain language statement:** Stress is common in nursing education and is often studied in terms of its negative effects. However, stressful experiences may also be viewed positively and contribute to motivation, confidence, learning, and personal growth. Few measures have been designed to capture this positive side of stress among nursing students.

We developed the Nursing Student Positive Stress Scale (NSPSS) to measure how nursing students positively appraise stressful experiences in both academic and clinical learning settings. The scale was informed by established stress theory and positive psychology. An international panel of nursing experts first reviewed the scale. It was then tested using responses from nursing students participating in a national survey in New Zealand.

The findings showed that the NSPSS performed well overall. The results supported related academic and clinical aspects of positive stress appraisal, and the scale showed good consistency across its items. Students who reported greater positive stress appraisal also tended to report lower levels of perceived stress.

The NSPSS provides a new way to examine positive responses to stress alongside existing measures that focus mainly on distress. It may help researchers and nursing educators develop a more balanced understanding of how students respond to challenging academic and clinical experiences. Further testing with nursing students in other settings is needed.

## Introduction

Stress remains a major concern within nursing education, as nursing students are required to manage academic expectations, clinical responsibilities, emotional demands, cultural awareness, and personal responsibilities while preparing for professional practice (Labrague, 2024). High stress levels have been associated with psychological distress, burnout, impaired wellbeing, academic difficulty, and programme withdrawal (Andargeery et al., 2024; Bakker et al., 2020), making students’ stress experiences important for designing educational environments that support mental health and professional development (Del Prato et al., 2011; Dias et al., 2024).

To date, measurement approaches in nursing education have primarily assessed stress stress exposure, perceived demands, and adverse responses (Labrague, 2024). The Perceived Stress Scale (PSS) is also widely used among nursing students to assess the extent to which life situations are perceived as unpredictable, uncontrollable, and overwhelming, although it is a generic measure and does not distinguish nursing-specific academic and clinical contexts (Cohen et al., 1983). The Student Nurse Stress Index (SNSI) a 22-item measure spanning academic load, clinical sources, interface worries, and personal problems, captures stress from both academic and clinical aspects of nursing education (Jones & Johnston, 1999). The Nursing Students’ Perceptions of Clinical Stressors Scale (NSPCSS) a 30-item instrument covering six domains including instructor conduct and clinical environment concerns, was developed specifically to identify stressors within the clinical environment (Rafati et al., 2021). Although these instruments provide context-specific assessments of nursing students’ stress, their measurements primarily focus on identifying stressors and the degree of perceived stress.

This predominant emphasis on stressors and adverse stress experiences provides an incomplete understanding of stress. The transactional theory of stress and coping proposes that stress responses are shaped by cognitive appraisal, whereby demands may be interpreted as threatening, harmful, or challenging depending on the perceived balance between demands and coping resources (Lazarus & Folkman, 1984). From this perspective, demanding experiences may not only contribute to distress, but may also be appraised as meaningful and manageable challenges when students perceive adequate support and resources (Gibbons et al., 2008, 2011; Lazarus & Folkman, 1984). This adaptive interpretation of stress is closely related to the concept of eustress, which refers to the positive outcome that results from stress experienced as beneficial (Selye, 1974). However, positive stress appraisal should not be understood simply as the absence of distress (Gibbons et al., 2008; Kloidt & Barsalou, 2024); it is the cognitive process of interpreting a demand as meaningful, manageable, and growth-promoting that is understood to give rise to eustress (Lazarus & Folkman, 1984). This concept is important in nursing education, where students are expected to develop competence, confidence, and professional identity through demanding learning experiences (Del Prato et al., 2011; Vabo et al., 2022). Understanding positive stress appraisal may therefore help explain how some educational demands support motivation, learning, and professional growth.

A small number of instruments have incorporated challenge appraisal or eustress more directly. The Clinical Stress Questionnaire (CSQ), an early application of cognitive appraisal theory to nursing, assesses whether students appraise clinical experiences as challenging or threatening, but is restricted to clinical experiences and conceptualises positive appraisal primarily as challenge (Pagana, 1989). The Index of Sources of Stress in Nursing Students (ISSN) extended this by explicitly incorporating both distress and eustress, asking students to rate course-related stressors as hassles or uplifts across learning and teaching, clinical placement, and course organisation domains; validation studies supported this distinction, with hassle ratings linked to poorer wellbeing and uplift ratings to healthier responses (Gibbons et al., 2008; Gibbons et al., 2009a, 2009b). Nevertheless, the ISSN primarily identifies which educational stressors function as hassles or uplifts, rather than directly measuring positive stress appraisal as across parallel academic and clinical contexts.

Beyond nursing education, several instruments further demonstrate that positive and negative aspects of stress can be measured as related but distinguishable constructs. The Valencia Eustress-Distress Appraisal Scale (VEDAS) assesses whether the same workplace demands are appraised as sources of eustress and distress across domains including workload, personal accountability, relationships, and home–work balance (Rodríguez et al., 2013). The Adolescent Distress-Eustress Scale (ADES) comprises separate five-item distress and eustress subscales and was developed specifically to capture both aspects of the stress response among adolescents (Branson et al., 2019). More recently, the Di-Eu-Stress State Scale (DESS) was developed to assess states of distress and eustress across undergraduate and employee samples, including ecological momentary assessment (Vikoler et al., 2025). These instruments provide important evidence that eustress and distress need not be conceptualised as opposite. However, they differ from the present measurement aim in population and context.

Overall, the available instruments reveal a specific gap. Nursing-specific measures predominantly assess stress sources or intensity, and although the CSQ and ISSN incorporate challenge appraisal or eustress, none assess positive stress appraisal using a structurally parallel academic and clinical format, despite these representing two major learning environments (Labrague, 2024; Pulido-Martos et al., 2012). Instruments developed outside nursing that distinguish eustress from distress, meanwhile, were designed for occupational, adolescent, or broader student and employee populations and do not reflect nursing’s particular educational demands. A nursing-specific instrument is therefore needed to assess positive stress appraisal consistently across academic and clinical contexts. The Nursing Student Positive Stress Scale (NSPSS) was developed to address this measurement gap.

## Aim

The aim of this study was to develop the NSPSS and evaluate its content validity, structural validity, internal consistency reliability, and convergent validity with perceived stress among nursing students.

## Methods

This methodological study formed Phase 2 of a sequential study investigating stress among nursing students in New Zealand. Phase 1 comprised a qualitative interview exploring nursing students’ perceptions and experiences of stress, findings from which informed the conceptual development of the NSPSS. Phase 2 developed and psychometrically evaluated the NSPSS, a tool designed to measure positive appraisal of academic and clinical stress experiences among nursing students. Scale development was guided by DeVellis’s scale development framework and involved theory-informed item development, expert content validation, and statistical evaluation of validity and reliability using data drawn from an ongoing national survey of nursing students in New Zealand (DeVellis, 2017). The study is reported in accordance with the Strengthening the Reporting of Observational Studies in Epidemiology (STROBE) guideline (von Elm et al., 2007).

### Item development

A deductive, theory-driven approach was used to develop the initial NSPSS item pool. The preceding qualitative phase identified that nursing students could perceive demanding academic and clinical experiences positively as well as negatively, supporting the relevance of positive stress appraisal within nursing education. Consistent with recommendations for instrument development in nursing education, the construct was theoretically defined and operationalised to guide item development (Mikkonen et al., 2022). Item development was guided by DeVellis’s scale development framework and informed by the transactional theory of stress and coping and positive psychology perspectives, particularly Fredrickson’s broaden-and-build theory. These theoretical perspectives were selected because the scale was designed to measure positive stress appraisal, rather than the presence of stressors or negative stress outcomes alone (Fredrickson, 2001; Lazarus & Folkman, 1984).

The transactional model provided the primary framework for conceptualising stress appraisal. Primary appraisal informed the challenge appraisal, personal growth, and motivation domains, reflecting that demands may be evaluated as opportunities rather than solely as threats; secondary appraisal informed the self-efficacy domain, concerning students’ perceived coping resources and confidence in managing high-pressure situations. This framework also supported assessing positive stress appraisal across the two major learning environments in which nursing students encounter different demands, academic and clinical (Lazarus & Folkman, 1984).

Fredrickson’s broaden-and-build theory provided the second theoretical foundation, informing the personal growth and satisfaction domains as potential positive consequences of successfully managing demanding experiences: personal growth reflects the perception that demanding experiences contribute to learning and development, while satisfaction captures the positive emotional response associated with successfully managing a challenge (Fredrickson, 2001; Fredrickson & Joiner, 2018).

Based on these theoretical foundations, five preliminary content domains were established: challenge appraisal, personal growth, motivation, self-efficacy, and satisfaction after overcoming challenges (Fredrickson, 2001; Lazarus & Folkman, 1984). Each domain was assessed across two contexts, academic learning and clinical learning, resulting in an initial 10-item scale consisting of five parallel item pairs. Items were rated from 1 (strongly disagree) to 5 (strongly agree), with higher summed scores indicating stronger positive stress appraisal; no items were reverse scored (Supplementary file).

### Content validity

Consistent with DeVellis’s scale development guidance, the initial item pool was reviewed by an expert panel to assess content validity before psychometric testing (DeVellis, 2017). Content validity was evaluated by an international panel of eight experts from New Zealand, Ireland, China and Thailand. All experts were registered nurses with clinical experience who also had experience in both nursing education and research. The expert panel included members with up to 40 years of higher nursing education experience.

Experts assessed item relevance using a four-point rating scale. The I-CVI was calculated based on relevance ratings as the proportion of experts rating an item as quite relevant or highly relevant. The S-CVI/Ave was calculated as the average of the I-CVI values across all items, while the S-CVI/UA was calculated as the proportion of items achieving universal agreement among experts (Lynn, 1986; Polit et al., 2007; Polit & Beck, 2006). Modified kappa statistics were also calculated to adjust I-CVI values for chance agreement. Following Lynn’s guidance, an I-CVI threshold of ≥ .78 was applied for a panel of six or more experts, and an S-CVI/Ave threshold of ≥ .90 was used to indicate adequate scale-level content validity. Expert comments, where provided, were reviewed before psychometric testing (Polit et al., 2007). Two experts (4 and 5) suggested wording revisions: concern that certain items implied stress was necessary for achievement, and suggestions to revise the retrospective phrasing of the satisfaction items. Both sets of original wording were retained, as the items were intended to assess stress as a potential contributor to motivation and performance rather than a necessary condition for it, and the satisfaction items’ retrospective framing was considered consistent with broaden-and-build theory’s emphasis on positive experiences building enduring personal resources (Fredrickson, 2001).

### Participants and data collection

Data for the psychometric evaluation were drawn from an ongoing national survey examining stress among nursing students in New Zealand, with responses collected between March and May 2026. Recruitment was conducted across thirteen tertiary education institutions in New Zealand, using convenience sampling through official institutional channels (email invitations and Moodle announcements) supplemented by snowball sampling; participation was voluntary. Eligible participants were students aged 18 years or older who were currently enrolled in undergraduate, postgraduate or enrolled nursing programmes in New Zealand. Participants completed demographic questions, the PSS-10 (Cohen et al., 1983), and the NSPSS. Data were collected using the Qualtrics online survey platform (Provo, UT, USA). A total of 539 responses were received during this period. Following the exclusion of one ineligible participant, one participant with missing NSPSS item data, and 30 multivariate outliers, 507 participants were included in the final psychometric analyses.

### Evaluation of validity and reliability

Data were screened before psychometric analysis. Missing data across the PSS-10 and NSPSS items were examined using Little’s MCAR test, and cases with missing scale data were excluded using listwise deletion. Multivariate outliers were assessed using Mahalanobis distance with a threshold of p < .001 (Finch, 2012; Little, 1988). Item distributions were examined using means, standard deviations, skewness, and kurtosis. For the PSS-10, items were scored from 0 (never) to 4 (very often). Positively worded items (4, 5, 7, and 8) were reverse scored before the item scores were summed, yielding a possible total score of 0–40, with higher scores indicating greater perceived stress (Cohen et al., 1983).

The analytical sample was randomly split into two non-overlapping subsamples by generating a uniformly distributed random number for each case in SPSS and assigning cases to groups using a cutoff of .50. This resulted in a subsample of 237 participants for exploratory factor analysis (EFA) and a separate subsample of 270 participants for confirmatory factor analysis (CFA) (Cabrera-Nguyen, 2010). The suitability of the EFA subsample for factor analysis was evaluated using the Kaiser-Meyer-Olkin (KMO) measure of sampling adequacy and Bartlett’s test of sphericity. A KMO value of ≥ .60 and a significant Bartlett’s test were considered to indicate adequate factorability (Dziuban & Shirkey, 1974; Kaiser, 1974).

EFA was conducted using principal axis factoring in the first subsample. Because the NSPSS was initially developed to reflect an overarching construct of positive stress appraisal, a one-factor solution was first examined (Costello & Osborne, 2005; Fabrigar et al., 1999). Rotation was not applied because only one factor was extracted. The number and adequacy of factors were evaluated using eigenvalues, scree plot inspection, communalities, factor loadings, and theoretical interpretability (Cattell, 1966; Costello & Osborne, 2005; Fabrigar et al., 1999). Cases with missing data were excluded listwise. Factor loadings with absolute values of ≥ .30 were considered acceptable (Watkins, 2018).

Multivariate normality was assessed in the Confirmatory factor analysis (CFA) subsample using Mardia’s multivariate skewness and kurtosis tests (Mardia, 1970). As the data departed from multivariate normality, CFA was conducted using robust maximum likelihood estimation (MLR), which provides robust standard errors and test statistics under non-normality (Curran et al., 1996; Rosseel, 2012). Model fit was evaluated using the chi-square statistic, comparative fit index (CFI), Tucker–Lewis index (TLI), root mean square error of approximation (RMSEA), and standardised root mean square residual (SRMR) (Schreiber et al., 2006). Conventional fit guidelines were used to support interpretation, with CFI and TLI values of ≥ .90 indicating acceptable fit and values of ≥ .95 indicating good fit. RMSEA and SRMR values of ≤ .08 were considered acceptable, with RMSEA values of ≤ .06 indicating good fit (Hu & Bentler, 1999).

Four theoretically informed CFA models were tested: a single-factor model (Model 1); the same model with residual covariances between corresponding academic and clinical item pairs added (Model 2); a two-correlated-factor model representing Academic and Clinical Positive Stress (Model 3); and the two-factor model with residual covariances added (Model 4), to account for potential shared wording and content overlap between parallel items(Schweizer et al., 2024).

Internal consistency reliability was evaluated using Cronbach’s alpha and McDonald’s omega (Cronbach, 1951; Dunn et al., 2014; McDonald, 1999). Reliability coefficients were calculated for the NSPSS total scale, the academic and clinical subscales, and the PSS-10. Corrected item–total correlations and alpha-if-item-deleted values were examined to evaluate item performance. Corrected item–total correlations of ≥ .30 were considered acceptable, and reliability coefficients of ≥ .70 were interpreted as acceptable for research purposes (Boateng et al., 2018; Nunnally & Bernstein, 1994).

Convergent validity was examined by testing the hypothesised relationship between NSPSS and PSS-10 scores. A small-to-moderate negative correlation was expected, as both measures concern subjective appraisal of stressful experiences but assess distinct aspects: the PSS-10 is a widely used, psychometrically validated measure of the extent to which life situations are appraised as stressful, unpredictable, and overwhelming (Cohen et al., 1983; Harris et al., 2023), whereas the NSPSS focuses specifically on positive appraisal of academic and clinical demands.

### Ethical considerations

Ethical approval was obtained from the University of Waikato Human Research Ethics Committee (HREC(Health)2025#57) on 24 July 2025. Institutional permission to distribute recruitment materials was obtained from relevant heads of school, programme managers, ethics committees, or designated research teams at participating tertiary institutions. Participants were provided with an online information sheet before entering the survey and indicated informed consent before participation. Participation was voluntary, and participants could choose not to answer any question or exit the survey before submission. Survey responses were collected anonymously and stored securely in accordance with university data management requirements. As the survey included questions about stress, information about student mental health services was provided to participants.

## Results

### Content validity

All ten NSPSS items met the predetermined content validity criteria. I-CVI values ranged from .88 to 1.00. Eight items achieved full expert agreement, with I-CVI values of 1.00. Two items, both relating to motivation in academic and clinical contexts, received agreement from seven of the eight experts and had I-CVI values of .88. These values were above the predetermined item-level threshold, and all ten items were retained for psychometric testing.

Modified kappa values ranged from .871 to 1.000, indicating excellent chance-corrected agreement across all items. At the scale level, the S-CVI/Ave was .975, exceeding the recommended threshold of .90, and the S-CVI/UA was .800, meeting the commonly used criterion of .80 for universal agreement (Polit et al., 2007; Polit & Beck, 2006). No item fell below the predetermined threshold, and all ten items were retained for psychometric testing.

### Data screening

A total of 539 survey responses were received. One response was excluded because the participant did not meet the eligibility criterion of being currently enrolled in a nursing programme, leaving 538 eligible responses for data screening. Missing data were examined across the PSS-10 and NSPSS items. Only one missing value was identified, occurring on the NSPSS clinical self-efficacy item, “I believe I can handle high-pressure situations”, representing 0.2% of eligible cases. Little’s MCAR test was non-significant, χ²(19) = 15.820, p = .669, indicating that the missing value was consistent with the assumption of missing completely at random (Little, 1988). Given the negligible level of missingness, listwise deletion was applied (Bennett, 2001).

After listwise deletion, 537 cases were available for multivariate outlier analysis. Mahalanobis distance was used to identify multivariate outliers across the scale items, using a conservative threshold of p < .001 (Finch, 2012). Thirty cases (5.6%) exceeded this threshold and were removed before psychometric analyses. The final analytical sample therefore consisted of 507 nursing students.

Item distributions were assessed before factor analysis and reliability testing to ensure that item responses showed sufficient variability and did not demonstrate severe departures from normality (Curran et al., 1996; Kim, 2013). Across the PSS-10 and NSPSS items, skewness values ranged from −0.971 to 0.093 and kurtosis values ranged from −0.842 to 1.001, indicating acceptable univariate normality.

### Characteristics of participants

The final analytical sample included 507 nursing students (Table 1). Of the 507 participants, most were female (88.4%, n = 448), and the largest age group was 20–25 years (40.0%, n = 203). Participants were enrolled across a range of nursing programmes, with the majority undertaking a General Bachelor of Nursing programme (70.8%, n = 359). Most participants were in Year 1 (34.5%, n = 175), Year 2 (35.3%, n = 179), or Year 3 (29.2%, n = 148). More than half reported part-time employment (51.3%, n = 260), and 26.4% reported full-time or part-time/shared caregiving responsibilities. Ethnicity was reported as a multi-select variable; the most frequently selected groups were New Zealand European/Pākehā (52.4%, n = 265), Asian (30.6%, n = 155), Māori (16.8%, n = 85), and Pasifika (8.5%, n = 43).

**Table 1:**
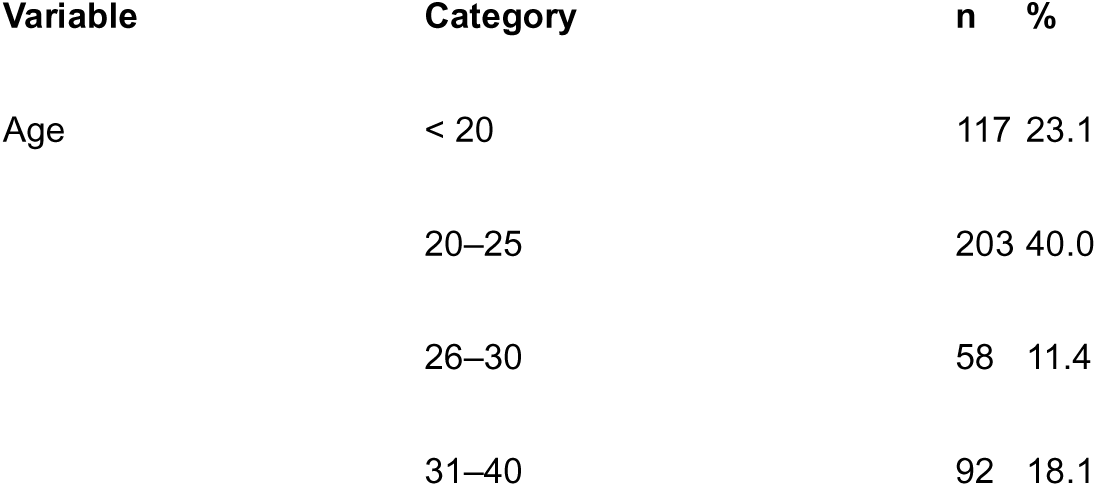
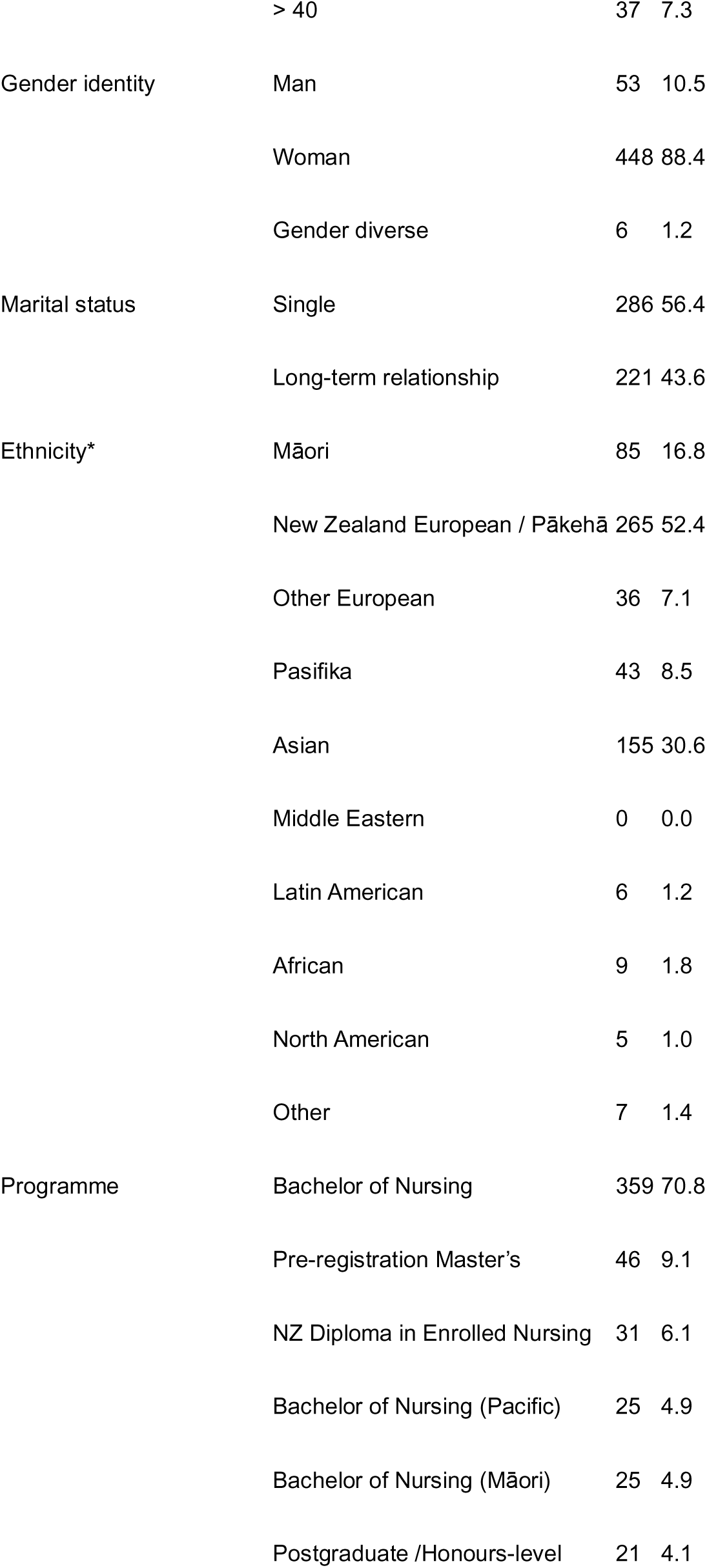
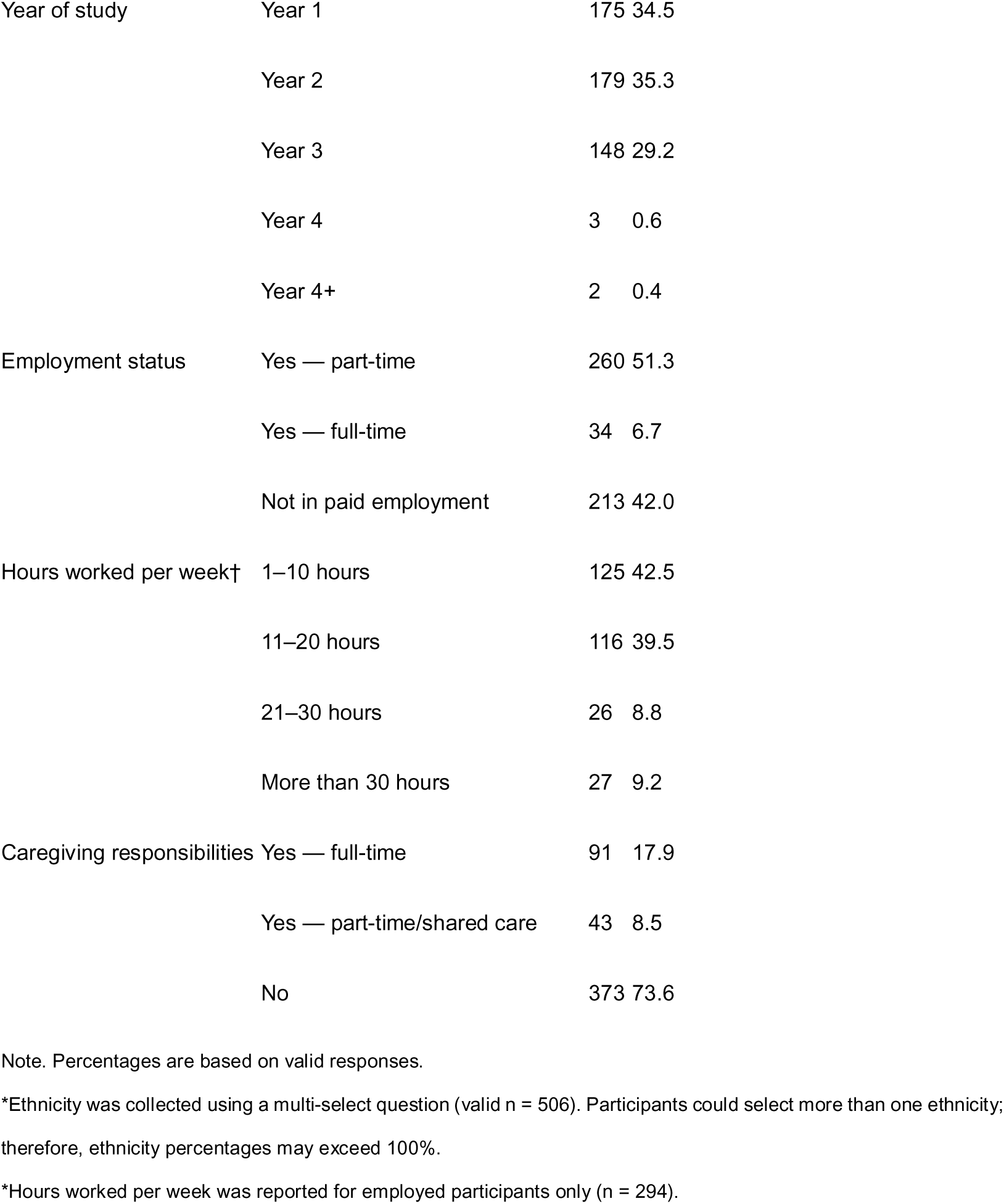
Demographic data.

### Exploratory factor analysis

EFA was conducted using the first randomly selected subsample (n = 237). The KMO value was .731, and Bartlett’s test of sphericity was significant, χ²(45) = 1342.23, p < .001, indicating that the correlation matrix was appropriate for factor analysis (Dziuban & Shirkey, 1974; Kaiser, 1974).

The NSPSS was developed to measure positive stress appraisal across academic and clinical contexts. Principal axis factoring was therefore conducted to examine whether the ten items reflected a general underlying factor. The first factor had an eigenvalue of 4.71, accounting for 47.09% of the total variance. Together with the theoretical conceptualisation of the scale, these findings supported a one-factor solution at this stage of analysis (Watkins, 2018).

All 10 items loaded positively on the extracted factor, with factor loadings ranging from .528 to .735 (Table 2). Most communalities were acceptable, although one item had an extraction communality slightly below .30. This item was retained because its factor loading was above .50 and it was conceptually consistent with the construct (Costello & Osborne, 2005; Hair et al., 2019).

**Table 2:** Exploratory Factor Analysis Item Statistics.

| Item | M (SD) | Factor loading | Communality |
| --- | --- | --- | --- |
| I view pressure as a positive challenge: Academic context | 3.30 (0.89) | .642 | .413 |
| I view pressure as a positive challenge: Clinical context | 3.42 (0.94) | .714 | .509 |
| Facing demands helps me grow: Academic context | 3.59 (0.87) | .724 | .524 |
| Facing demands helps me grow: Clinical context | 3.75 (0.85) | .735 | .540 |
| Stress pushes me to perform at my best: Academic context | 2.88 (1.12) | .624 | .389 |
| Stress pushes me to perform at my best: Clinical context | 2.89 (1.17) | .646 | .418 |
| I believe I can handle high-pressure situations: Academic context | 3.54 (0.91) | .625 | .391 |
| I believe I can handle high-pressure situations: Clinical context | 3.65 (0.89) | .528 | .279 |
| I feel satisfied after overcoming challenges: Academic context | 4.21 (0.83) | .596 | .355 |
| I feel satisfied after overcoming challenges: Clinical context | 4.29 (0.81) | .566 | .321 |

### Confirmatory factor analysis

Multivariate normality of the 10 NSPSS items was assessed on the CFA subsample (n = 270) using Mardia’s test. Both multivariate skewness (b1p = 11.98, χ² = 539.04, p < .001) and multivariate kurtosis (b2p = 140.71, z = 10.98, p < .001) indicated significant departure from multivariate normality. CFA models were therefore estimated using MLR. Although EFA supported a single dominant factor consistent with a coherent underlying construct of positive stress appraisal, the NSPSS was designed with parallel item pairs across academic and clinical contexts. CFA was therefore used to test whether this broader construct was better represented as a single unidimensional factor or as two correlated, context-specific factors, given the scale’s dual-context design. The four models specified a priori (see Methods) were tested in sequence.

The initial unidimensional model showed poor fit to the data, χ²(35) = 587.49, p < .001, CFI = .480, TLI = .332, RMSEA = .242, and SRMR = .134. Adding correlated residuals between the five academic and clinical versions of the same items improved model fit but remained inadequate, χ²(30) = 271.72, p < .001, CFI = .773, TLI = .659, RMSEA = .173, and SRMR = .173. A two-factor model distinguishing academic and clinical positive stress also showed poor fit without correlated residuals, χ²(34) = 536.30, p < .001, CFI = .528, TLI = .375, RMSEA = .234, and SRMR = .129.

The final model specified two correlated factors, Academic Positive Stress and Clinical Positive Stress, with correlated residuals between the five parallel academic and clinical item pairs. This model demonstrated good fit on CFI and TLI and acceptable fit on RMSEA, χ²(29) = 60.49, p = .001, CFI = .970, TLI = .954, RMSEA = .063, 90% CI [.041, .086], and SRMR = .065. The model was therefore retained as the best-fitting CFA solution.

Standardised factor loadings were statistically significant (p < .001), with loadings ranging from .416 to .717 for the Academic Positive Stress factor and from .372 to .675 for the Clinical Positive Stress factor (Table 3). The two latent factors were moderately correlated (r = .542), consistent with the interpretation that academic and clinical positive stress appraisal represent related, context-specific dimensions of the broader construct. The specified residual correlations between same-wording academic and clinical item pairs were also statistically significant (p < .001), with estimates ranging from .420 to .799, consistent with shared item-specific variance associated with the parallel item format (Bandalos, 2021; Brown, 2015; Schweizer et al., 2024).

**Table 3:** Standardised estimates for the final model.

| Parameter | Estimate | SE | z | p |
| --- | --- | --- | --- | --- |
| Academic Positive Stress |  |  |  |  |
| Q15_1: View pressure as challenge | .700 | .053 | 13.317 | < .001 |
| Q16_1: Demands help me grow | .717 | .045 | 15.985 | < .001 |
| Q17_1: Stress pushes me to perform at my best | .603 | .050 | 12.079 | < .001 |
| Q18_1: Handle high-pressure situations | .574 | .048 | 12.046 | < .001 |
| Q19_1: Satisfied after overcoming challenges | .416 | .058 | 7.211 | < .001 |
| Clinical Positive Stress |  |  |  |  |
| Q15_2: View pressure as challenge | .675 | .055 | 12.276 | < .001 |
| Q16_2: Demands help me grow | .647 | .059 | 10.897 | < .001 |
| Q17_2: Stress pushes me to perform at my best | .673 | .052 | 12.898 | < .001 |
| Q18_2: Handle high-pressure situations | .489 | .059 | 8.276 | < .001 |
| Q19_2: Satisfied after overcoming challenges | .372 | .057 | 6.513 | < .001 |
| Correlated residuals between same-wording item pairs |  |  |  |  |
| Q15_1 ~~ Q15_2 | .420 | .072 | 5.860 | < .001 |
| Q16_1 ~~ Q16_2 | .551 | .070 | 7.840 | < .001 |
| Q17_1 ~~ Q17_2 | .431 | .069 | 6.291 | < .001 |
| Q18_1 ~~ Q18_2 | .547 | .063 | 8.712 | < .001 |
| Q19_1 ~~ Q19_2 | .799 | .034 | 23.334 | < .001 |

*Note.* Estimates are standardised. The retained model specified two correlated latent factors, Academic Positive Stress and Clinical Positive Stress, with correlated residuals between parallel academic and clinical versions of the same item.

### Reliability

Internal consistency reliability was assessed using Cronbach’s alpha (α) and McDonald’s omega (ω) in the full analytical sample (N = 507) (Cronbach, 1951; Dunn et al., 2014). The NSPSS 10-item full scale demonstrated good internal consistency (α = .839, ω = .843), and two 5-item context-specific subscales showed acceptable reliability: Academic Positive Stress (range 5-25, M= 17.39, SD= 3.28; α = .765 and ω = .774), Clinical Positive Stress (range 5-25, M= 17.91, SD= 3.19; α = .743 and ω = .751). Corrected item–total correlations exceeded the recommended .30 threshold throughout with full scale .431-.636; Academic .434-.633; Clinical .342-.602, supporting item retention (Boateng et al., 2018). The PSS-10 comparison measure (range 0-40, M= 22.08, SD= 5.79) similarly demonstrated good internal consistency (α = .841, ω = .843, item-total correlations .413-.702). The close correspondence between alpha and omega across measures indicated consistent reliability estimates (Dunn et al., 2014).

### Convergent validity

Convergent validity was examined by evaluating the relationship between the NSPSS total score and the PSS-10 total score. A significant negative correlation was observed, r = −.298, p < .001, N = 507. This indicated a modest inverse association between positive stress appraisal and perceived stress, suggesting that the two constructs are related but not overlapping (Terwee et al., 2007).

## Discussion

This study developed and psychometrically evaluated the NSPSS as a measure of positive stress appraisal among nursing students, providing preliminary evidence across content validity, structural validity, internal consistency reliability, and convergent validity.

The EFA identified a dominant general positive stress appraisal factor, with all ten items loading positively on the extracted factor. However, this unidimensional structure was not supported in the subsequent CFA. CFA also did not support a simple two-factor structure distinguishing academic and clinical positive stress.

Acceptable model fit was achieved only when two contextual factors and residual dependencies between all five parallel academic and clinical item pairs were modelled simultaneously. Therefore, the findings support a two-context measurement model incorporating item-specific dependencies arising from the parallel item design, rather than a simple one-or two-factor structure.

The two latent factors may reflect context-specific expressions of a broader positive stress appraisal construct. Their moderate correlation (r = .542) suggests that academic and clinical positive stress appraisals are related but not identical. This distinction is meaningful in nursing education, where academic demands commonly include examinations, assignments, knowledge acquisition, and performance expectations, whereas clinical demands may involve patient care responsibilities, professional socialisation, communication with clinical staff, and managing uncertainty in practice settings (Pulido-Martos et al., 2012). As anticipated, acceptable model fit required specifying correlated residuals between the five parallel academic and clinical item pairs, consistent with the a priori expectation that identical item wording presented across two contexts would generate shared item-specific variance independent of the substantive academic-clinical distinction (Bandalos, 2021; Schweizer et al., 2024). The relatively high residual correlation between the two satisfaction items (r = .799) indicates particularly strong shared item-specific variance beyond that explained by the academic and clinical factors, which may reflect the closely related emotional content of the two satisfaction items relative to the other four item pairs. Replication in an independent sample is recommended to confirm that this two-factor, method-effect-adjusted structure remains stable, and to establish whether the magnitude of specific residual correlations, particularly the satisfaction item pair, is consistent across samples.

The NSPSS demonstrated good internal consistency reliability for the full scale and acceptable reliability for the two context-specific item sets: Academic Positive Stress and Clinical Positive Stress. The close correspondence between Cronbach’s alpha and McDonald’s omega estimates suggested that the reliability findings were consistent across coefficients (Dunn et al., 2014). Given that each item set contains only five items, these acceptable reliability estimates support their preliminary use in examining academic and clinical expressions of positive stress appraisal (Cortina, 1993; Tavakol & Dennick, 2011). The total score may be used provisionally to represent overall positive stress appraisal, while the academic and clinical scores may provide context-specific information. Further research is needed to determine the most appropriate scoring structure.

A negative correlation was observed between the NSPSS and PSS-10. This association is theoretically interpretable because both measures concern students’ subjective appraisal of stressful experiences (Cohen et al., 1983; Lazarus & Folkman, 1984). The modest magnitude of this association provides preliminary empirical support for conceptualising positive stress appraisal as related to, but distinguishable from, perceived stress. Whereas the PSS-10 assesses the extent to which life demands are experienced as unpredictable, uncontrollable, and overwhelming, the NSPSS captures positive interpretations of demanding academic and clinical experiences, including challenge, growth, motivation, coping confidence, and satisfaction after overcoming difficulties (Cohen et al., 1983). Students may therefore experience educational demands as stressful while simultaneously perceiving aspects of those experiences as meaningful or growth-promoting. This coexistence of positive and negative responses has also been identified in qualitative research. Nursing students have described academic and clinical experiences as sources of both distress and eustress (Gibbons et al., 2008),while graduate nursing students experiencing the combined demands of research and clinical practicum have reported psychological strain alongside adjustment and personal and professional growth (An et al., 2026). Similar findings among international university students indicate that stressful experiences may involve both distress and eustress, depending on how demands are perceived and managed (Gong & Geertshuis, 2023).

As outlined earlier, existing nursing-specific measures (e.g., ISSN, SNSI) assess stress sources or general eustress rather than positive appraisal specifically. The present findings extend this literature by demonstrating that positive appraisal can be reliably measured as a parallel academic–clinical construct, with the two context factors showing a moderate but non-redundant correlation (r = .542). This structural distinction, not previously tested in existing instruments, supports treating academic and clinical positive appraisal as related but separable, rather than assuming a single general eustress factor in nursing education.

### Limitation

Several limitations should be considered. First, convenience sampling and voluntary participation may have introduced selection and self-selection bias, as students more motivated or interested in stress may have been more likely to participate, limiting generalisability; future studies should use more representative recruitment strategies and, where population data are available, consider weighting procedures. Second, EFA and CFA were conducted using subsamples from the same dataset, so validation in an independent sample is needed to assess the stability and replicability of the factor structure, including the five correlated residuals in the final CFA model, across cohorts and educational contexts. Finally, convergent validity was assessed using the PSS-10, an indirect comparator because it measures perceived stress rather than eustress or positive appraisal; future studies should examine additional validity evidence, including measurement invariance, predictive validity, and associations with outcomes such as coping, resilience, engagement, and wellbeing.

### Implications

The NSPSS has implications for both nursing education research and practice. As a measure of positive stress appraisal, it may be used alongside existing stress and wellbeing measures to provide a more balanced understanding of nursing students’ stress experiences. Rather than focusing only on stressors or distress-related outcomes, the scale enables researchers and educators to examine how students interpret demanding academic and clinical experiences in terms of challenge, growth, motivation, coping confidence, and satisfaction after overcoming difficulties (Gibbons et al., 2008; Lazarus & Folkman, 1984).

In practice, the NSPSS may help identify context-specific patterns in students’ positive appraisal of stress across academic and clinical learning environments. This may be useful for understanding how students respond to different types of educational demands, such as academic workload, assessment pressure, clinical placement expectations, and uncertainty in practice settings. Such information could inform learning support strategies that not only reduce harmful stress but also strengthen students’ capacity to appraise manageable challenges as meaningful and growth-promoting (Del Prato et al., 2011; Rudland et al., 2020). The NSPSS may therefore complement distress-focused assessments and support a more approach to student mental health and wellbeing in nursing education.

## Conclusion

This study developed and provided initial validation evidence for the NSPSS, a theory-informed measure of positive stress appraisal spanning challenge appraisal, personal growth, motivation, coping confidence, and satisfaction across academic and clinical learning contexts. By focusing on how students positively interpret demanding educational experiences, the NSPSS offers a complementary perspective to existing stressor-and distress-focused measures and may contribute to a more balanced understanding of nursing students’ stress experiences in future research and educational practice.

## Declaration of Interest Statement

The authors declare no competing interests.

## Funding statement

This research did not receive any specific grant from funding agencies in the public, commercial, or not-for-profit sectors.

## Supporting information

Supplementary File 1: STROBE checklist

Supplementary File 2: Nursing Student Positive Stress Scale (NSPSS)

## Data Availability

The data that support the findings of this study are available from the corresponding author upon reasonable request, subject to ethical and privacy considerations.

## Acknowledgment

The author would like to thank the participants and institutions for their input.

## Notes

### Competing Interest Statement

The authors have declared no competing interest.

### Author Declarations

The University of Waikato Human Research Ethics Committee gave ethical approval for this work HREC(Health)2025#57.

## References

1. An, R., Cheng, J., Lan, X., Han, X., Liao, W., Lin, L., Chen, M., & Zhang, Y. (2026). Psychological adjustment and growth of graduate nursing students under the dual stress of research and practicum: A qualitative study. Frontiers in Medicine, 13. 10.3389/fmed.2026.1734998

2. Andargeery, S. Y., Taani, M. H., Alhalwani, R. A., & El-Gazar, H. E. (2024). Psychological Distress, Academic Stress, and Burnout among Saudi Undergraduate Nursing Students. Journal of Clinical Medicine, 13(12), 3357. 10.3390/jcm13123357

3. Bakker, E. J. M., Kox, J. H. A. M., Boot, C. R. L., Francke, A. L., van der Beek, A. J., & Roelofs, P. D. D. M. (2020). Improving mental health of student and novice nurses to prevent dropout: A systematic review. Journal of Advanced Nursing, 76(10), 2494–2509. 10.1111/jan.14453

4. Bandalos, D. L. (2021). Item Meaning and Order as Causes of Correlated Residuals in Confirmatory Factor Analysis. Structural Equation Modeling: A Multidisciplinary Journal, 28(6), 903–913. 10.1080/10705511.2021.1916395

5. Bennett, D. A. (2001). How can I deal with missing data in my study? Australian and New Zealand journal of public health, 25(5), 464–469.

6. Boateng, G. O., Neilands, T. B., Frongillo, E. A., Melgar-Quiñonez, H. R., & Young, S. L. (2018). Best Practices for Developing and Validating Scales for Health, Social, and Behavioral Research: A Primer. Frontiers in Public Health, Volume 6-2018. 10.3389/fpubh.2018.00149

7. Branson, V., Dry, M. J., Palmer, E., & Turnbull, D. (2019). The Adolescent Distress-Eustress Scale: Development and validation. SAGE Open, 9(3), 1–14. 10.1177/2158244019865802

8. Brown, T. A. (2015). Confirmatory Factor Analysis for Applied Research, Second Edition. Guilford Publications.

9. Cabrera-Nguyen, P. (2010). Author Guidelines for Reporting Scale Development and Validation Results in the Journal of the Society for Social Work and Research. Journal of the Society for Social Work and Research, 1(2), 99–103. JSTOR. 10.5243/jsswr.2010.8

10. Cattell, R. B. (1966). The Scree Test For The Number Of Factors. Multivariate Behavioral Research, 1(2), 245–276. 10.1207/s15327906mbr0102_10

11. Cohen, S., Kamarck, T., & Mermelstein, R. (1983). A Global Measure of Perceived Stress. Journal of Health and Social Behavior, 24(4), 385–396. JSTOR. 10.2307/2136404

12. Cortina, J. M. (1993). What is coefficient alpha? An examination of theory and applications. Journal of Applied Psychology, 78(1), 98–104. 10.1037/0021-9010.78.1.98

13. Costello, A. B., & Osborne, J. (2005). Best practices in exploratory factor analysis: Four recommendations for getting the most from your analysis. 10.7275/JYJ1-4868

14. Cronbach, L. J. (1951). Coefficient alpha and the internal structure of tests. Psychometrika, 16(3), 297–334. 10.1007/BF02310555

15. Curran, P. J., West, S. G., & Finch, J. F. (1996). The robustness of test statistics to nonnormality and specification error in confirmatory factor analysis. Psychological Methods, 1(1), 16–29. 10.1037/1082-989X.1.1.16

16. Del Prato, D., Bankert, E., Grust, P., & Joseph, J. (2011). Transforming nursing education: A review of stressors and strategies that support students’ professional socialization. Advances in Medical Education and Practice, 2, 109–116. 10.2147/AMEP.S18359

17. DeVellis, R. F. (2017). Scale development: Theory and applications (4th ed.). SAGE Publications.

18. Dias, J. M., Subu, M. A., Al-Yateem, N., Ahmed, F. R., Rahman, S. A., Abraham, M. S., Forootan, S. M., Sarkhosh, F. A., & Javanbakh, F. (2024). Nursing students’ stressors and coping strategies during their first clinical training: A qualitative study in the United Arab Emirates. BMC Nursing, 23(1), 322. 10.1186/s12912-024-01962-5

19. Dunn, T. J., Baguley, T., & Brunsden, V. (2014). From alpha to omega: A practical solution to the pervasive problem of internal consistency estimation. British Journal of Psychology, 105(3), 399–412. 10.1111/bjop.12046

20. Dziuban, C. D., & Shirkey, E. C. (1974). When is a correlation matrix appropriate for factor analysis? Some decision rules. Psychological Bulletin, 81(6), 358–361. 10.1037/h0036316

21. Fabrigar, L. R., Wegener, D. T., MacCallum, R. C., & Strahan, E. J. (1999). Evaluating the Use of Exploratory Factor Analysis in Psychological Research.

22. Finch, W. H. (2012). Distribution of Variables by Method of Outlier Detection. Frontiers in Psychology, 3, 211. 10.3389/fpsyg.2012.00211

23. Fredrickson, B. L. (2001). The Role of Positive Emotions in Positive Psychology. The American Psychologist, 56(3), 218–226. 10.1037//0003-066x.56.3.218

24. Fredrickson, B. L., & Joiner, T. (2018). Reflections on Positive Emotions and Upward Spirals. Perspectives on Psychological Science, 13(2), 194–199. 10.1177/1745691617692106

25. Gibbons, C., Dempster, M., & Moutray, M. (2008). Stress and eustress in nursing students. Journal of Advanced Nursing, 61(3), 282–290. 10.1111/j.1365-2648.2007.04497.x

26. Gibbons, C., Dempster, M., & Moutray, M. (2009a). Index of sources of stress in nursing students: a confirmatory factor analysis. Journal of advanced nursing, 65(5), 1095–1102.

27. Gibbons, C., Dempster, M., & Moutray, M. (2009b). Surveying nursing students on their sources of stress: A validation study. Nurse Education Today, 29(8), 867–872.

28. Gibbons, C., Dempster, M., & Moutray, M. (2011). Stress, coping and satisfaction in nursing students. Journal of Advanced Nursing, 67(3), 621–632. 10.1111/j.1365-2648.2010.05495.x

29. Gong, W., & Geertshuis, S. A. (2023). Distress and eustress: An analysis of the stress experiences of offshore international students. Frontiers in Psychology, 14. 10.3389/fpsyg.2023.1144767

30. Hair, J. F., Black, W. C., Babin, B. J., & Anderson, R. E. (2019). Multivariate Data Analysis. Cengage.

31. Harris, K. M., Gaffey, A. E., Schwartz, J. E., Krantz, D. S., & Burg, M. M. (2023). The Perceived Stress Scale as a Measure of Stress: Decomposing Score Variance in Longitudinal Behavioral Medicine Studies. Annals of Behavioral Medicine: A Publication of the Society of Behavioral Medicine, 57(10), 846–854. 10.1093/abm/kaad015

32. Hu, L., & Bentler, P. M. (1999). Cutoff criteria for fit indexes in covariance structure analysis: Conventional criteria versus new alternatives. Structural Equation Modeling: A Multidisciplinary Journal, 6(1), 1–55. 10.1080/10705519909540118

33. Jones, M. C., & Johnston, D. W. (1999). The derivation of a brief Student Nurse Stress Index. Work & Stress, 13(2), 162–181. 10.1080/026783799296129

34. Kaiser, H. F. (1974). An index of factorial simplicity. Psychometrika, 39(1), 31–36. 10.1007/BF02291575

35. Kim, H.-Y. (2013). Statistical notes for clinical researchers: Assessing normal distribution (2) using skewness and kurtosis. Restorative Dentistry & Endodontics, 38(1), 52–54. 10.5395/rde.2013.38.1.52

36. Kloidt, J., & Barsalou, L. W. (2024). Establishing a Comprehensive Hierarchical construct of Eustress (CHE). Current Psychology (New Brunswick, N.j.), 43(41), 32258–32273. 10.1007/s12144-024-06750-7

37. Labrague, L. J. (2024). Umbrella Review: Stress Levels, Sources of Stress, and Coping Mechanisms among Student Nurses. Nursing Reports, 14(1), 362–375. 10.3390/nursrep14010028

38. Lazarus, R. S., & Folkman, S. (1984). Stress, Appraisal, and Coping. Springer Publishing Company. https://books.google.com.au/books?id=i-ySQQuUpr8C

39. Little, R. J. A. (1988). A Test of Missing Completely at Random for Multivariate Data with Missing Values. Journal of the American Statistical Association, 83(404), 1198–1202. 10.1080/01621459.1988.10478722

40. Lynn, M. R. (1986). Determination and quantification of content validity. Nursing Research, 35(6), 382–385.

41. Mardia, K. V. (1970). Measures of Multivariate Skewness and Kurtosis with Applications. Biometrika, 57(3), 519–530. 10.2307/2334770

42. McDonald, R. P. (1999). Test Theory: A Unified Treatment. L. Erlbaum Associates.

43. Mikkonen, K., Tomietto, M., & Watson, R. (2022). Instrument development and psychometric testing in nursing education research. Nurse education today, 119, 105603. 10.1016/j.nedt.2022.105603

44. Nunnally, J. C., & Bernstein, I. H. (1994). Psychometric Theory. McGraw-Hill Companies,Incorporated.

45. Pagana Kathleen Deska. (1989). Psychometric Evaluation of the Clinical Stress Questionnaire (CSQ). Journal of Nursing Education, 28(4), 169–174. 10.3928/0148-4834-19890401-07

46. Polit, D. F., & Beck, C. T. (2006). The content validity index: Are you sure you know what’s being reported? Critique and recommendations. Research in Nursing & Health, 29(5), 489–497. 10.1002/nur.20147

47. Polit, D. F., Beck, C. T., & Owen, S. V. (2007). Is the CVI an acceptable indicator of content validity? Appraisal and recommendations. Research in Nursing & Health, 30(4), 459–467. 10.1002/nur.20199

48. Pulido-Martos, M., Augusto-Landa, J. m., & Lopez-Zafra, E. (2012). Sources of stress in nursing students: A systematic review of quantitative studies. International Nursing Review, 59(1), 15–25. 10.1111/j.1466-7657.2011.00939.x

49. Rafati, F., Sharif Nia, H., Khoshnood, Z., & Allen, K.-A. (2021). Development and psychometric testing of nursing students’ perceptions of clinical stressors scale: An instrument design study. BMC Psychiatry, 21(1), 1. 10.1186/s12888-020-02964-8

50. Rodríguez, I., Kozusznik, M. W., & Peiró, J. M. (2013). Development and validation of the Valencia Eustress-Distress Appraisal Scale. International Journal of Stress Management, 20(4), 279.

51. Rosseel, Y. (2012). lavaan: An R Package for Structural Equation Modeling. Journal of Statistical Software, 48, 1–36. 10.18637/jss.v048.i02

52. Rudland, J. R., Golding, C., & Wilkinson, T. J. (2020). The stress paradox: How stress can be good for learning. Medical Education, 54(1), 40–45. 10.1111/medu.13830

53. Schreiber, J. B., Nora, A., Stage, F. K., Barlow, E. A., & King, J. (2006). Reporting Structural Equation Modeling and Confirmatory Factor Analysis Results: A Review. The Journal of Educational Research, 99(6), 323–338. 10.3200/JOER.99.6.323-338

54. Schweizer, K., Gold, A., Krampen, D., & Troche, S. (2024). Conceptualizing Correlated Residuals as Item-Level Method Effects in Confirmatory Factor Analysis. Educational and Psychological Measurement, 84(5), 869–886. 10.1177/00131644231218401

55. Selye, H. (1974). Stress without distress. In Psychopathology of human adaptation (pp. 137-146). Springer.

56. Tavakol, M., & Dennick, R. (2011). Making sense of Cronbach’s alpha. International Journal of Medical Education, 2, 53–55. 10.5116/ijme.4dfb.8dfd

57. Terwee, C. B., Bot, S. D. M., De Boer, M. R., Van Der Windt, D. A. W. M., Knol, D. L., Dekker, J., Bouter, L. M., & De Vet, H. C. W. (2007). Quality criteria were proposed for measurement properties of health status questionnaires. Journal of Clinical Epidemiology, 60(1), 34–42. 10.1016/j.jclinepi.2006.03.012

58. Vabo, G., Slettebø, Å., & Fossum, M. (2022). Nursing students’ professional identity development: An integrative review. Nordic Journal of Nursing Research, 42(2), 62–75. 10.1177/20571585211029857

59. Vikoler, T., Kovács, D., & Traut-Mattausch, E. (2025). The Di-Eu-Stress State Scale (DESS Scale): Development and validation of a scale measuring state distress and eustress. European Journal of Psychological Assessment, 41(6), 486.

60. von Elm, E., Altman, D. G., Egger, M., Pocock, S. J., Gøtzsche, P. C., & Vandenbroucke, J. P. (2007). The Strengthening the Reporting of Observational Studies in Epidemiology (STROBE) Statement: Guidelines for reporting observational studies. PLoS Medicine, 4(10), e296. 10.1371/journal.pmed.0040296

61. Watkins, M. W. (2018). Exploratory Factor Analysis: A Guide to Best Practice. Journal of Black Psychology, 44(3), 219–246. 10.1177/0095798418771807

