## Supplementary File 1: STROBE checklist for "Measuring Positive Stress Appraisal Among Nursing Students: Development and Psychometric Evaluation of the Nursing Student Positive Stress Scale (NSPSS)"

STROBE Statement—checklist of items that should be included in reports of observational studies

|  | Item No. | Recommendation | Page  No. | Relevant text from manuscript |
| --- | --- | --- | --- | --- |
| **Title and abstract** | 1 | (*a*) Indicate the study’s design with a commonly used term in the title or the abstract | 1 | Title: "Measuring Positive Stress Appraisal Among Nursing Students in New Zealand: Development and Psychometric Evaluation of the Nursing Student Positive Stress Scale (NSPSS)"; Abstract Design line: "A methodological instrument development and psychometric evaluation study." |
|  |  | (*b*) Provide in the abstract an informative and balanced summary of what was done and what was found | 1 | Structured abstract: Background, Objective, Design, Methods, Results, Conclusion |
| Introduction | | | |  |
| Background/rationale | 2 | Explain the scientific background and rationale for the investigation being reported | 3-5 | “Stress remains a major concern within nursing education, as nursing students are required to manage academic expectations…” |
| Objectives | 3 | State specific objectives, including any prespecified hypotheses | 5 | "This study aimed to evaluate its content validity, structural validity, internal consistency reliability..." |
| Methods | | | |  |
| Study design | 4 | Present key elements of study design early in the paper | 6 | "This methodological study formed Phase 2 of a sequential study investigating stress among nursing students in New Zealand..." |
| Setting | 5 | Describe the setting, locations, and relevant dates, including periods of recruitment, exposure, follow-up, and data collection | 8 | "Data for the psychometric evaluation were drawn from an ongoing national survey examining stress among nursing students..." |
| Participants | 6 | (*a*) *Cohort study*—Give the eligibility criteria, and the sources and methods of selection of participants. Describe methods of follow-up  *Case-control study*—Give the eligibility criteria, and the sources and methods of case ascertainment and control selection. Give the rationale for the choice of cases and controls  *Cross-sectional study*—Give the eligibility criteria, and the sources and methods of selection of participants | 8 | "Eligible participants were students aged 18 years or older who were currently ..." Convenience sampling via institutional channels (email, Moodle) supplemented by snowball sampling; participation voluntary. |
|  |  | (*b*) *Cohort study*—For matched studies, give matching criteria and number of exposed and unexposed  *Case-control study*—For matched studies, give matching criteria and the number of controls per case |  | Not applicable: cross-sectional design without matching. |
| Variables | 7 | Clearly define all outcomes, exposures, predictors, potential confounders, and effect modifiers. Give diagnostic criteria, if applicable | 6-9 | Five theory-derived domains (challenge appraisal, personal growth, motivation, self-efficacy, satisfaction), each rated across Academic and Clinical contexts, 1-5 Likert. Participants and data collection / Evaluation of validity and reliability (p.8-9): PSS-10 (Cohen et al., 1983) scored 0-4, items 4/5/7/8 reverse scored, total range 0-40. |
| Data sources/ measurement | 8* | For each variable of interest, give sources of data and details of methods of assessment (measurement). Describe comparability of assessment methods if there is more than one group | 6-9 | NSPSS developed using DeVellis's scale development framework, informed by transactional theory of stress and coping and Fredrickson's broaden-and-build theory; content validity assessed by an 8-member international expert panel; data collected via Qualtrics online survey platform alongside PSS-10 and demographic items. |
| Bias | 9 | Describe any efforts to address potential sources of bias | 9, 17 | Evaluation of validity and reliability: Mahalanobis distance (p<.001) used to identify multivariate outliers; Little's MCAR test used to assess missingness mechanism; robust MLR estimator used in CFA given non-normality. |
| Study size | 10 | Explain how the study size was arrived at | 8-9 | "A total of 539 responses were received during this period... 507 participants were included in the final psychometric analyses"; analytical sample randomly split into EFA subsample (n=237) and CFA subsample (n=270). |

Continued on next page

| Quantitative variables | 11 | Explain how quantitative variables were handled in the analyses. If applicable, describe which groupings were chosen and why | 6, 9 | NSPSS items rated on a 1 (strongly disagree) to 5 (strongly agree) Likert scale, summed with no reverse scoring. Analytical sample randomly split (uniform random number, cutoff .50) into non-overlapping EFA (n=237) and CFA (n=270) subsamples. |
| --- | --- | --- | --- | --- |
| Statistical methods | 12 | (*a*) Describe all statistical methods, including those used to control for confounding | 9-10 | EFA: principal axis factoring, KMO/Bartlett's test for factorability, eigenvalues/scree plot/loadings for factor retention. CFA: robust maximum likelihood (MLR) estimation; fit evaluated via chi-square, CFI, TLI, RMSEA, SRMR. Reliability: Cronbach's alpha and McDonald's omega. Convergent validity: correlation between NSPSS and PSS-10 total scores. |
|  |  | (*b*) Describe any methods used to examine subgroups and interactions |  | Not applicable: no subgroup or interaction analyses were conducted. |
|  |  | (*c*) Explain how missing data were addressed | 9, 11 | "Missing data across the PSS-10 and NSPSS items were examined using Little's MCAR test, and cases with missing scale data were excluded using listwise deletion". |
|  |  | (*d*) *Cohort study*—If applicable, explain how loss to follow-up was addressed  *Case-control study*—If applicable, explain how matching of cases and controls was addressed  *Cross-sectional study*—If applicable, describe analytical methods taking account of sampling strategy | 8-9 | Convenience sampling through institutional channels supplemented by snowball sampling; random split of the analytic sample into EFA and CFA subsamples described. |
|  |  | (*e*) Describe any sensitivity analyses | 13-14 | No separate sensitivity analyses; instead four a priori CFA models (unidimensional with/without correlated residuals; two-factor with/without correlated residuals) were compared to test competing structural hypotheses (Confirmatory factor analysis). |
| Results | | | | |
| Participants | 13* | (a) Report numbers of individuals at each stage of study—eg numbers potentially eligible, examined for eligibility, confirmed eligible, included in the study, completing follow-up, and analysed | 11-12 | "A total of 539 survey responses were received. One response was excluded because the participant did not meet the eligibility criterion... The final analytical sample therefore consisted of 507 nursing students" |
|  |  | (b) Give reasons for non-participation at each stage |  | Not ascertainable. Recruitment was conducted through institutional distribution and an anonymous online survey; therefore, the number and reasons for eligible students who did not participate were unavailable. |
|  |  | (c) Consider use of a flow diagram |  | N/A |
| Descriptive data | 14* | (a) Give characteristics of study participants (eg demographic, clinical, social) and information on exposures and potential confounders | 12 | Characteristics of participants (Table 2) |
|  |  | (b) Indicate number of participants with missing data for each variable of interest | 11 | "Only one missing value was identified, occurring on the NSPSS clinical self-efficacy item... representing 0.2% of eligible cases". |
|  |  | (c) *Cohort study*—Summarise follow-up time (eg, average and total amount) |  | N/A: cross-sectional design, no follow-up period. |
| Outcome data | 15* | *Cohort study*—Report numbers of outcome events or summary measures over time |  | N/A |
|  |  | *Case-control study—*Report numbers in each exposure category, or summary measures of exposure |  | N/A |
|  |  | *Cross-sectional study—*Report numbers of outcome events or summary measures | 12-15 | EFA factor loadings and variance explained; CFA fit indices for the final model; Cronbach's alpha/McDonald's omega for NSPSS and subscales; NSPSS-PSS-10 correlation. |
| Main results | 16 | (*a*) Give unadjusted estimates and, if applicable, confounder-adjusted estimates and their precision (eg, 95% confidence interval). Make clear which confounders were adjusted for and why they were included | 12-15 | EFA: dominant factor, eigenvalue=4.71, 47.09% variance explained, loadings .528-.735 (p.12-13). CFA final model: χ²(29)=60.49, p=.001, CFI=.970, TLI=.954, RMSEA=.063 [90% CI .041-.086], SRMR=.065 (p.13-14). Reliability: NSPSS α=.839, ω=.843; Academic subscale α=.765, ω=.774; Clinical subscale α=.743, ω=.751; PSS-10 α=.841, ω=.843 (p.14). Convergent validity: r=-.298, p<.001, N=507. No confounder adjustment was applicable to this psychometric evaluation. |
|  |  | (*b*) Report category boundaries when continuous variables were categorized |  | N/A: no continuous variables were categorised in the analyses. |
|  |  | (*c*) If relevant, consider translating estimates of relative risk into absolute risk for a meaningful time period |  | N/A |

Continued on next page

| Other analyses | 17 | Report other analyses done—eg analyses of subgroups and interactions, and sensitivity analyses | 13-14 | Not a subgroup/sensitivity analysis in the epidemiological sense; four a priori competing CFA models were tested and compared to identify the best-fitting structural model. |
| --- | --- | --- | --- | --- |
| Discussion | | | | |
| Key results | 18 | Summarise key results with reference to study objectives | 15-16 | "This study developed and psychometrically evaluated the NSPSS…The findings provided evidence of strong content validity, preliminary structural validity, preliminary convergent validity... and good internal consistency reliability". |
| Limitations | 19 | Discuss limitations of the study, taking into account sources of potential bias or imprecision. Discuss both direction and magnitude of any potential bias | 17 | Limitations: convenience/voluntary sampling and potential selection/self-selection bias; EFA and CFA conducted on subsamples from the same dataset, requiring replication in an independent sample; correlated residuals in the final CFA model require replication testing; PSS-10 as an indirect convergent validity comparator. |
| Interpretation | 20 | Give a cautious overall interpretation of results considering objectives, limitations, multiplicity of analyses, results from similar studies, and other relevant evidence | 15-17 | Discussion: interpretation of the two-factor, method-effect-adjusted CFA structure; moderate correlation between Academic and Clinical latent factors (r=.542) interpreted as related but distinct context-specific dimensions; negative NSPSS-PSS-10 correlation interpreted as evidence that positive stress appraisal and perceived stress are related but distinguishable constructs. |
| Generalisability | 21 | Discuss the generalisability (external validity) of the study results | 17-18 | "This limits the generalisability of the findings. Future studies should use more representative recruitment strategies...". |
| Other information | |  | | |
| Funding | 22 | Give the source of funding and the role of the funders for the present study and, if applicable, for the original study on which the present article is based | 19 | "This research did not receive any specific grant from funding agencies in the public, commercial, or not-for-profit sectors." |

*Give information separately for cases and controls in case-control studies and, if applicable, for exposed and unexposed groups in cohort and cross-sectional studies.

**Note:** An Explanation and Elaboration article discusses each checklist item and gives methodological background and published examples of transparent reporting. The STROBE checklist is best used in conjunction with this article (freely available on the Web sites of PLoS Medicine at http://www.plosmedicine.org/, Annals of Internal Medicine at http://www.annals.org/, and Epidemiology at http://www.epidem.com/). Information on the STROBE Initiative is available at www.strobe-statement.org.
