## Supplementary File 2: Nursing Student Positive Stress Scale (NSPSS) for "Measuring Positive Stress Appraisal Among Nursing Students: Development and Psychometric Evaluation of the Nursing Student Positive Stress Scale (NSPSS)"

Please indicate how strongly you agree with each statement.

1 = Strongly disagree	2 = Disagree	3 = Neutral	4 = Agree	5 = Strongly agree

| Item | Context | 1 | 2 | 3 | 4 | 5 |
| --- | --- | --- | --- | --- | --- | --- |
| 1. I view pressure as a positive challenge | Academic | ☐ | ☐ | ☐ | ☐ | ☐ |
|  | Clinical | ☐ | ☐ | ☐ | ☐ | ☐ |
| 2. Facing demands helps me grow | Academic | ☐ | ☐ | ☐ | ☐ | ☐ |
|  | Clinical | ☐ | ☐ | ☐ | ☐ | ☐ |
| 3. Stress pushes me to perform at my best | Academic | ☐ | ☐ | ☐ | ☐ | ☐ |
|  | Clinical | ☐ | ☐ | ☐ | ☐ | ☐ |
| 4. I believe I can handle high-pressure situations | Academic | ☐ | ☐ | ☐ | ☐ | ☐ |
|  | Clinical | ☐ | ☐ | ☐ | ☐ | ☐ |
| 5. I feel satisfied after overcoming challenges | Academic | ☐ | ☐ | ☐ | ☐ | ☐ |
|  | Clinical | ☐ | ☐ | ☐ | ☐ | ☐ |
